# Steps on the path to clinical translation – a survey of the UK MRI community

**DOI:** 10.1101/2024.12.20.24319456

**Authors:** Julia E Markus, Penny L Hubbard Cristinacce, Shonit Punwani, James PB O’Connor, Rebecca Mills, Maria Yanez Lopez, Matthew Grech-Sollars, Fabrizio Fasano, John C Waterton, Michael J Thrippleton, Matt G Hall, Susan T Francis, Ben Statton, Kevin Murphy, Po-Wah So, Harpreet Hyare

## Abstract

**Purpose:** Our goal was to understand the barriers and challenges to clinical translation of quantitative MR (qMR) as perceived by stakeholders un the UK.

**Methods:** We conducted an electronic survey on 7 key areas related to clinical translation of qMR, developed at the BIC-ISMRM workshop: “Steps on the path to clinical translation”. Based on the 7 areas identified: (i) clinical workflow, (ii) changes in clinical practice, (ii) improving validation, (iv) standardisation of data acquisition and analysis, (v) sharing of data and code, (vi) improving quality management, and (vii) end-user engagement, a 40-question survey was developed. Members of BIC-ISMRM, MR-PHYSICS, BSNR and institutional mailing lists were invited to respond to the online survey over a 5-week period between September and October 2022. The responses were analysed via descriptive statistics of multiple-choice questions, Likert scores and a thematic analysis of free text questions.

**Results:** A total of 69 responses were received from predominantly research imaging scientists (69%) in numerous centres across the UK. Three main themes were identified: 1) the need to develop Consensus in terminology, decision making and validation; 2) an appreciation of uniqueness of each clinical situation and the need for Context Dependency; and 3) the importance of developing a Product Profile as early in development as possible.

**Conclusion:** Effective translation of qMR imaging and spectroscopic biomarkers to achieve their full clinical in the UK and internationally must address the differing needs and expectations of a wide range of stakeholders.

## 1. Introduction

Quantitative magnetic resonance (qMR) imaging and spectroscopic biomarkers can probe a multitude of biophysical properties in patients with a wide range of diseases. They offer great potential for advancing our understanding of pathology as well as improving diagnosis, prognosis and prediction of response to therapy^1^. Too often, this potential has not translated into widespread clinical adoption, with a low number of qMR imaging or spectroscopic methods used in clinical decision-making^2^. A workshop was held in September 2022 to highlight to the British and Irish Chapter of the International Society for Magnetic Resonance in Medicine (BIC-ISMRM) community the difficulties in translating qMR imaging and spectroscopic biomarkers, developed in academia, into clinical applications and to discuss some possible solutions to improve successful translation^3^.

Workshop attendees heard the perspectives of several Invited speakers: a radiologist, a radiographer, clinical physicists, a vendor, an imaging Contract/Clinical Research Organizations (CROs), an open science networks, a metrologist, an imaging network, and those developing consensus methods. A round-table discussion was held in which workshop participants discussed a range of questions relevant to clinical translation of qMR imaging and spectroscopic biomarkers. Each of the seven groups summarised their findings via three main conclusions and three further questions. These questions were used as the basis of the online survey of the broader UK MR community presented here.

Previous surveys of the MRI community have involved obtaining information about the use of MRI in areas such as radiotherapy^4^ and neuroradiology^5^, and in the use of specific methods, such as Perfusion MRI^6^. Surveys have also assessed the use of quality assurance and quality control^7^programmes for MRI, of the geographical availability of MRI scanners^8^. Many of these aspects are of relevance to the clinical translation of qMR imaging and spectroscopic biomarkers, but in general, these surveys were exercises in fact-finding and did not canvas the opinions of the community. This survey aims to ascertain the opinions of the UK MRI community on qMR clinical translation in 7 key areas: clinical workflow, changes in clinical practice, improving validation, standardisation of data acquisition and analysis, sharing of data and code, improving quality management, and end-user engagement.

## 2. Methods

### 2.1 Survey

On 7^th^ September 2022, the BIC-ISMRM held a workshop entitled “Steps on the path to clinical translation” in Cardiff. Following a series of talks by a range of stakeholders involved in the clinical translation of qMR biomarkers, participants were allocated into small groups to discuss one of seven questions related to the translation of MR imaging biomarkers. Participants were asked to provide three main conclusions and three further questions arising from these group-facilitated discussions and presented these at the subsequent panel discussion. Further details of the workshop can be found in a previous publication^3^. The questions produced from these discussions were used to form a survey aimed at capturing the opinions and knowledge of the wider UK MR community. Where possible the survey authors adhered to the questions derived via consensus during the roundtable discussion. However, some changes to the wording and format were made for the purposes of clarity and ease of analysis.

A REDCap project was used to derive a web-based e-survey a QR/quick access link distributed as an open invitation. The survey was open for 5 weeks, between 9^th^ September to 17^th^ October 2022. It was made available during the 2022 BIC-ISMRM Annual Meeting which followed the workshop and subsequently distributed via communication email lists and newsletters to the BIC-ISMRM, MR-PHYSICS, British Society of Neuroradiologists (BSNR) and various institutional emailing lists. The survey was composed of 40 questions covering 7 topics (Table 1, column 1).

**Table 1.** Survey questions (column 1) generated from the questions posed at the roundtable discussions at the BIC-ISMRM workshop. Response type for each question is given in column 2 along with the percentage of participants who gave that answer. Free text answers for each question can be found in Table S1 of the supplementary material. Survey found: https://redcap.slms.ucl.ac.uk/surveys/index.php?s=5PdwR6JJdLvK5SfT. Question 3: Data are presented for those in a clinical role (clinicians, radiographers and imaging scientists in a clinical role) and those in a research role (imaging scientist in a research role). Those describing their role as “other” were not included.

| Survey questions | Answers |
| --- | --- |
| **Q1**<br>A. Is an imaging biomarker useful if it doesn't give a yes/no answer to aid diagnosis?<br>B. How can we standardise the language we use to aid clinical translation? Should effort be made to develop a consensus paper to standardise terminology to aid clinical translation?<br>C. Quantitative MR parameter maps often need to be created away from the scanner console. What is the maximum amount of time after acquisition, if an imaging biomarker is to be integrated into the clinical workflow? | A. No (5%), Yes (83%), Unsure (9%)<br>B. Free text (**Table S1**) and No (3%), Yes (69%), Unsure (28%)<br>C. a) The subject still needs to be on the scanner (23%)<br>b) Within 1 hour of acquisition (13%)<br>c) Within 24 hours of acquisition (44%)<br>d) Before the subject's next visit (20%) |
| **Q2**<br>A. In regards to sensitivity and specificity, what percentage of change is significant enough to change practice? How should this be defined?<br>B. Who should be involved in the decision to change practice?<br>C. Should change in clinical practice be for all patients or selected cohorts? | A. a) 1-2% (7%); b) 2-10% (14%); c) 10-30% (53%); d) 30-50% (12%) e) over 50% (14%) and free text (**Table S1**).<br>B. Multiple answers possible a) Clinicians (97%); b) Hospital Managers (62%); c) Radiographers (71%); d) Physicists (75%); e) Other (23%)<br>C. a) All Patients (8%); b) Selected Cohorts (37%); c) Unsure (55%) |
| **Q3**<br>A. Does "validation" mean something different to a physicist and a clinician?<br>How much do you agree with the following statements.<br>i. "Validation is knowing an imaging biomarker's repeatability and reproducibility."<br>ii. "Validation is knowing the accuracy and precision of the measurement."<br>iii. "Validation is making a clinician happy to use your imaging biomarker." | Ai. a) Strongly Agree (22%); b) Agree (46%); c) Neither agree or disagree (4%); d) Disagree (9%); e) Strongly Disagree (3%)<br>Aii. a) Strongly Agree (14%); b) Agree (48%); c) Neither agree or disagree (9%); d) Disagree (10%); e) Strongly Disagree (1%)<br>Aiii. a) Strongly Agree (4%); b) Agree (22%); c) Neither agree or disagree (14%); d) Disagree (30%); e) Strongly Disagree (13%)<br>Aiv. a) Strongly Agree (19%); b) Agree (33%); c) Neither agree or |
| iv. "Validation is knowing how well we can measure an imaging biomarker."<br>B. At what point do you consider something validated?<br>C. How do we incentivise transparency and reproducibility?<br>How much do you agree with the following statement.<br>i. "Funders need to mandate sharing of code."<br>ii. "Funders need to mandate sharing of data."<br>iii. "Publishers need to mandate sharing of code."<br>iv. "Publishers need to mandate sharing of data."<br>v. "Institutions need to provide knowledge support for code sharing."<br>vi. "Institutions need to provide knowledge support for data sharing." | disagree (22%); d) Disagree (9%); e) Strongly Disagree (3%)<br>B. Free text (*Table S1*).<br>Ci. a) Strongly Agree (23%); b) Agree (43%); c) Neither agree or disagree (21%); d) Disagree (9%); e) Strongly Disagree (4%)<br>Cii. a) Strongly Agree (12%); b) Agree (44%); c) Neither agree or disagree (28%); d) Disagree (11%); e) Strongly Disagree (5%)<br>Ciii. a) Strongly Agree (28%); b) Agree (26%); c) Neither agree or disagree (26%); d) Disagree (14%); e) Strongly Disagree (5%)<br>Civ. a) Strongly Agree (14%); b) Agree (33%); c) Neither agree or disagree (32%); d) Disagree (14%); e) Strongly Disagree (7%)<br>Cv. a) Strongly Agree (50%); b) Agree (41%); c) Neither agree or disagree (7%); d) Disagree (2%); e) Strongly Disagree (0%)<br>Cvi. a) Strongly Agree (53%); b) Agree (35%); c) Neither agree or disagree (11%); d) Disagree (2%); e) Strongly Disagree (0%) |
| **Q4**<br>A. It is known that vendors will find a method less attractive if a method requires a phantom, so when should we use a phantom? Should we use phantoms to standardise data acquisition for every sequence?<br>B. Do you agree with a goal-orientated approach to quality?<br>C. Should we standardise, harmonise or optimise pulse sequences in multi-centre studies? | A. Free text (*Table S1*) and No (48%) /Yes (5%) /Unsure (43%)<br>B. No (0%) /Yes (52%) /Unsure (48%)<br>C. a) Standardise (24%); b) Harmonise (42%); c) Optimise (11%); d) Unsure (24%) |
| **Q5** | Ai. a) From the very beginning of a project (2%); b) At the point |
| **A. i. At what point should we be sharing code?**<br>**ii. At what point should we be sharing data?**<br>**B. How do we balance the need for the subject's privacy against the value of sharing data? i.e. how much can we anonymise without losing valuable information and how do we get consent for making data public?**<br>**C. Techniques generally require buy-in from scanner manufacturers to become part of their product in order to change clinical practice. How do we balance the need for researchers to protect the IP to allow this to happen, against the value of sharing code and data publicly?** | of publication (**73%**); c) Once the method is established (**25%**); d) Never (**0%**)<br>Aii. a) From the very beginning of a project (**5%**); b) At the point of publication (**71%**); c) Once the method is established (**18%**) d) Never (**5%**)<br>B. Free text (*Table S1*)<br>C. Free text (*Table S1*) |
| **Q6**<br>**A. Does a Quality Management System (QMS) exist in your place of work?**<br>**B. Do you use it?**<br>**C. How do we incentivise the use of a QMS?** | A. No (**20%**), Yes (**46%**), Unsure (**33%**)<br>B. No (**50%**), Yes (**34%**), Unsure (**16%**)<br>C. Free text (*Table S1*) |
| **Q7**<br>**A. As an early career researcher, what's the best way to contact clinicians?**<br>**B. How do we integrate our MR development with PACs systems?**<br>**C. How do we find out if a patient group can tolerate the imaging method?** | A. Free text (*Table S1*)<br>B. Free text (*Table S1*)<br>C. Free text (*Table S1*) |

### 2.2 Descriptive analysis

Basic occupational demographic questions allowed us to monitor MRI community representation. These included: Role in Institution, Institution Type, and Area of Imaging Research. Descriptive statistics were drawn from multiple choice (MCQ), Likert (a rating scale that asks respondents to indicate their level of agreement or disagreement with a statement or question) and agree/disagree questions.

The percentage of survey participants who agreed with a response was calculated for each question for all of those who replied to that question. For questions 2B, 3B and 3C, further analysis allowed the descriptive statistics to be calculated for sub-groups of participants, as described in the results.

### 3.2 Thematic analysis

Thematic analysis was utilised as the technique of choice to analyse all free-text responses.

Founded in psychology research, thematic analysis, is a structured yet flexible method of performing data analysis in qualitative research. By following a pre-determined set of steps or phases commonalities across data sets can be identified, organised and assigned meaning to the research question^9^. Thematic Analysis has been adopted across the healthcare landscape ^10,11,12^ as a useful methodology for researchers to develop core skills in qualitative analysis^10,13,14^.

Thematic analysis with a deductive approach^9^ was used and tested on the whole of the free text data against a specific research question. The steps we used are shown in Figure 1 and described below:

1) **Decide on the format in which to obtain the qualitative data** — free-form comment boxes allowed unstructured responses from survey participants. 14 of the sub-questions were free-text responses, and each of the 7 sections had additional space for free-text comments. This allowed responses to be included outside of the direct question (see Supplementary Material).
2) **Establish a basis for a deductive approach** — our hypothetical ground truth was that qMR sequences can translate to clinical practice with a defined pipeline that builds enough practical information to be used clinically from a concerted effort by research collaborators, clinical workforce, and patients.
3) **Become familiar with the responses** — Responses were read and re-read multiple times and notes taken on possible common responses.
4) **Apply codes to the responses** — Common responses were identified by codes.
5) **Generating themes** — A concept or label was developed that gave a meaningful description to the initial codes.
6) **Reviewing themes** — Transcripts were re-read with these themes in mind and the appropriateness assessed.
7) **Defining themes** — Themes were defined based on the cohesion of the codes within the themes and boundaries redrawn where appropriate.
8) **Naming themes** — Themes were named based on specific and unique characteristics.

**Figure 1.**
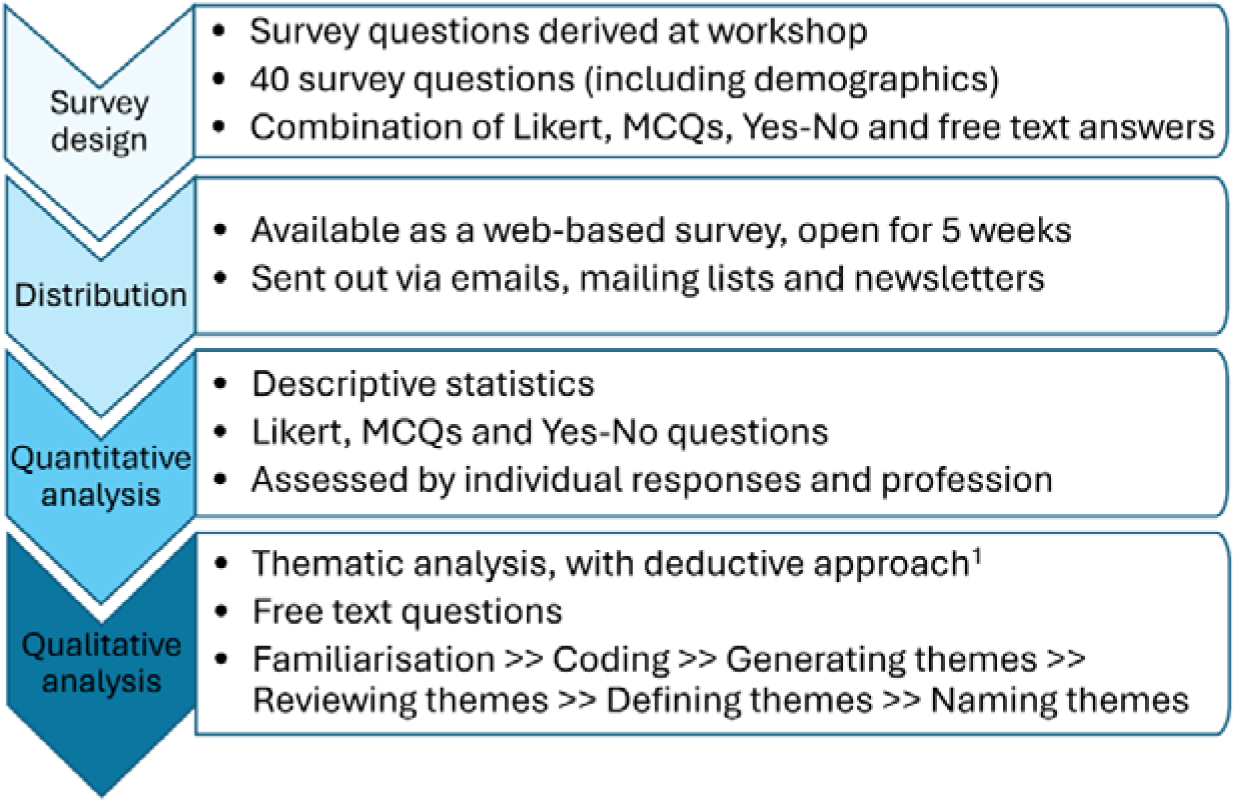
Schematic of the analysis of the results of the UK-based survey on the clinical translation of qMR imaging and spectroscopic biomarkers.

## 3. Results

### 3.1 Demographics

We received 69 responses with the completion rate varying from 41–100% of the survey. Responses were received from Imaging Scientists (Research (64%) and Clinical (18%)), Clinicians (10%), and other (8%). The type of institution in which the participants work was primarily Universities (56%), followed by NHS hospital trusts (27%), Private clinics/Independent sector(7%) and finally other (10%). The area of imaging research was most commonly Neuroimaging (64%), followed by Cancer Imaging (42%), Cardiac Imaging (14%), Musculoskeletal (14%) and other (16%), with many participants active in more than one area.

### 3.2 Descriptive analysis

Table 1 details for each of the 7 sections the questions asked in the survey (column 1) and details the percentage of participants who answered each question (column 2).

Section 1 was developed from the question **‘What will and won’t work in a clinical workflow?’** and provided three sub-questions for the survey. 83% of participants agreed that an imaging biomarker is still useful if it does not give a yes/no answer to aid diagnosis (Q1A) and 69% agreed that effort should be made to develop a consensus paper to standardise terminology to aid clinical translation (Q1B). Responses were more variable for Q1C, ‘Quantitative MR parameter maps often need to be created away from the scanner console. What is the maximum amount of time after acquisition, if an imaging biomarker is to be integrated into the clinical workflow?’. 44% stated ‘within 24 hours of acquisition’, 23% ‘whilst the subject still needs to the on the scanner’, 20% ‘before the subject’s next visit’, and 13% said ‘within 1 hour of acquisition’.

Section 2 was based on the roundtable discussion question **‘How big of an improvement justifies a change in clinical practice?** Q2A asked, ‘In regard to sensitivity and specificity, what percentage of change is significant enough to change practice?’. Although 53% selected 10-30% change as significant enough to change practice, there was similar distribution amongst the remaining answers. When asked to give a multi-choice answer to ‘Who should be involved in the decision to change practice?’ (Q2B) almost everyone agreed that clinicians should be involved (97%), followed by Physicists (75%), Radiographers (71%) and Hospital Managers (62%). When comparing the answers from those in a Clinical Role (i.e. Clinicians, Clinical Imaging Scientists and Radiographers) to those in a Research Role (Research Imaging Scientists) we find that there was broad agreement regarding who should be involved, except regarding the role of Hospital Managers (Figure 2). There was a high degree of uncertainty when asked ‘Should change in clinical practice be for all patients or selected cohorts?’. 55% said “unsure” and 37% “for selected cohorts”.

**Figure 2.**
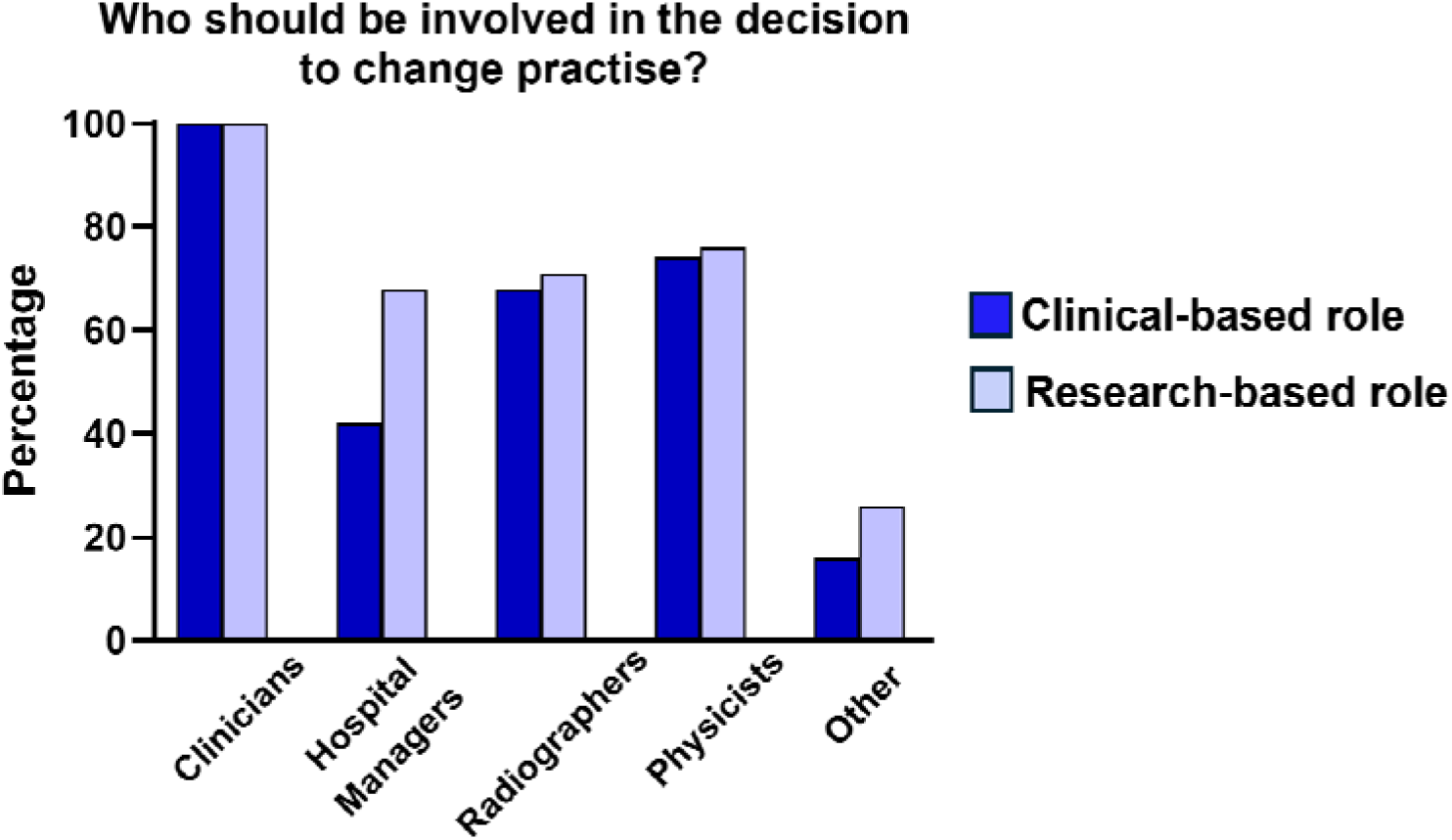
Comparison of the percentage of those in Clinical and Research Roles when ask ‘Who should be involved in the decision to change practice?’ (Q2B).

Section 3 questions were developed from the question **‘How can we improve validation of an imaging biomarker?’** Firstly, Q3A asked ‘Does “validation” mean something different to a physicist and a clinician?’ and gave four definitions for participants to agree/disagree with. Figure 3 shows the results, divided into the Clinical Role v Research Role groups. Similar trends are seen, although the strength of agreement was generally stronger for those in a Clinical Role. The strongest agreement was that “Validation is knowing an imaging biomarker’s repeatability and reproducibility” (68% of all 69 participants strongly agreed or agreed) and the weakest agreement was that “Validation is making a clinician happy to use your imaging biomarker” (26% of all 69 participants strongly agreed or agreed).

**Figure 3.**
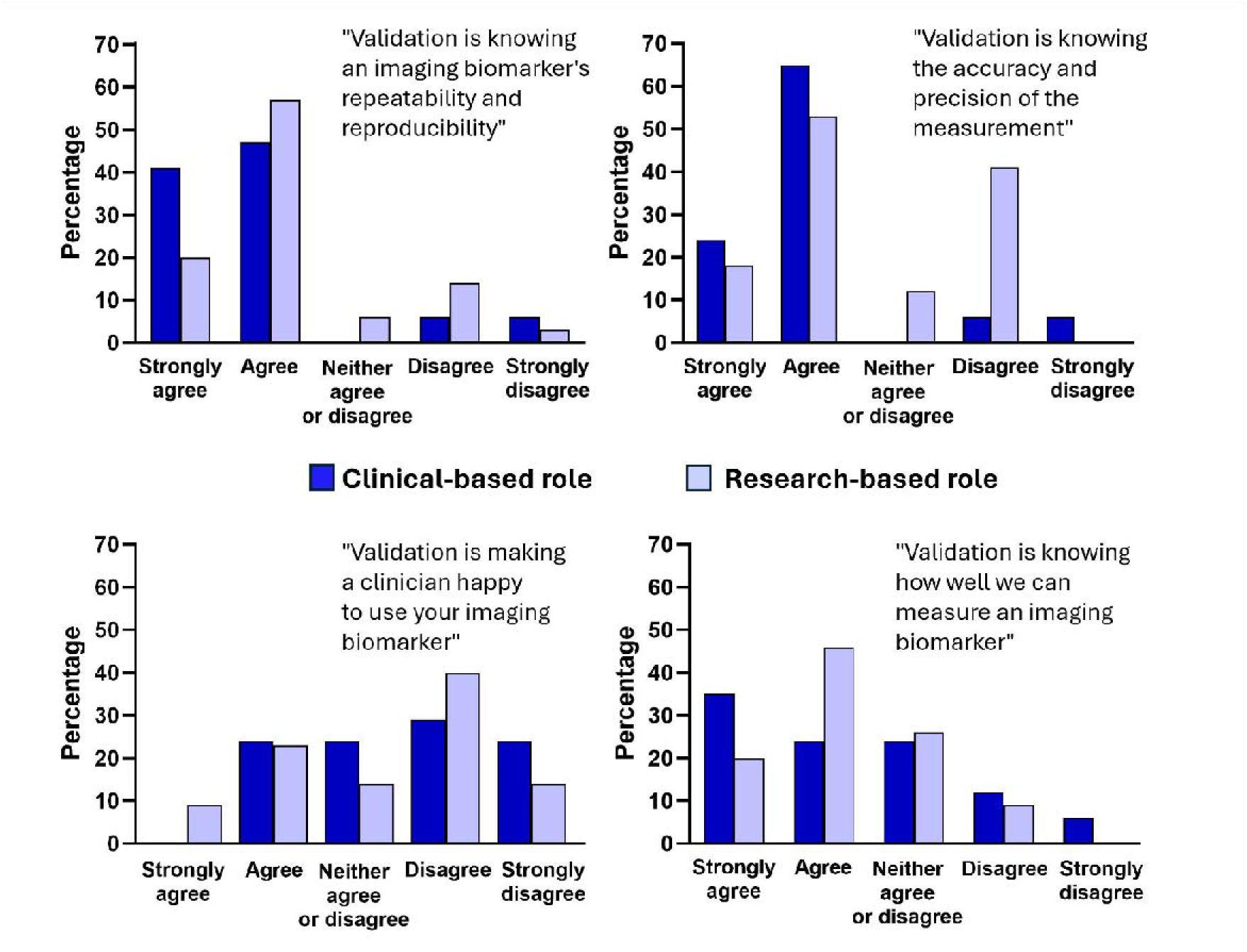
Q3A asked the question, ‘Does validation mean something different to a physicist and a clinician?’. Four definitions of validation were proposed, and participants were asked to say if they agreed (on a 5-point scale). Data are presented for those in a Clinical Role (clinicians, radiographers and imaging scientists in a clinical role) and those in a Research Role (imaging scientist in a research role). Those describing their role as “other” were not included.

Q3C asked ‘How do we incentivise transparency and reproducibility?’ and gave participants the chance to agree or disagree, again on a 5-point scale. Most agreed/strongly agreed that code sharing should be mandated by both funders (66%) and publishers (54%), but results were less clear for data (funders: 56%, and publishers: 47%). However, there was a high consensus that institutions should provide knowledge support for data (88%) and code (91%) sharing (Figure 4).

**Figure 4.**
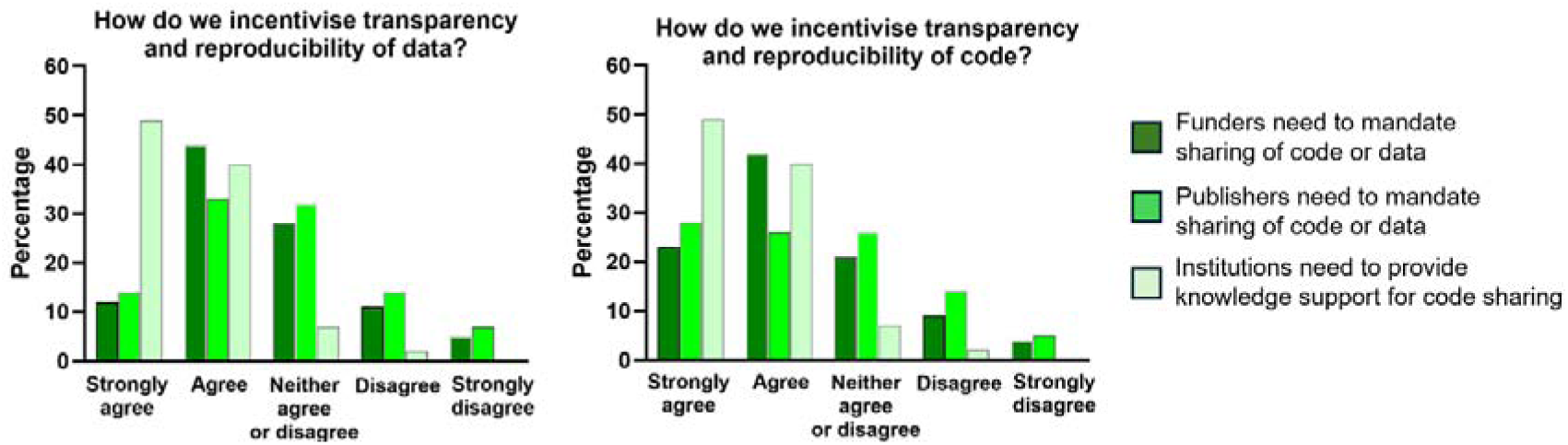
Strength of agreement (on a 5-point scale) that funders and publishers should mandate sharing and that institutions should provide knowledge support for, (a) data sharing, and (b) code sharing (Q3C).

**Figure 5.**
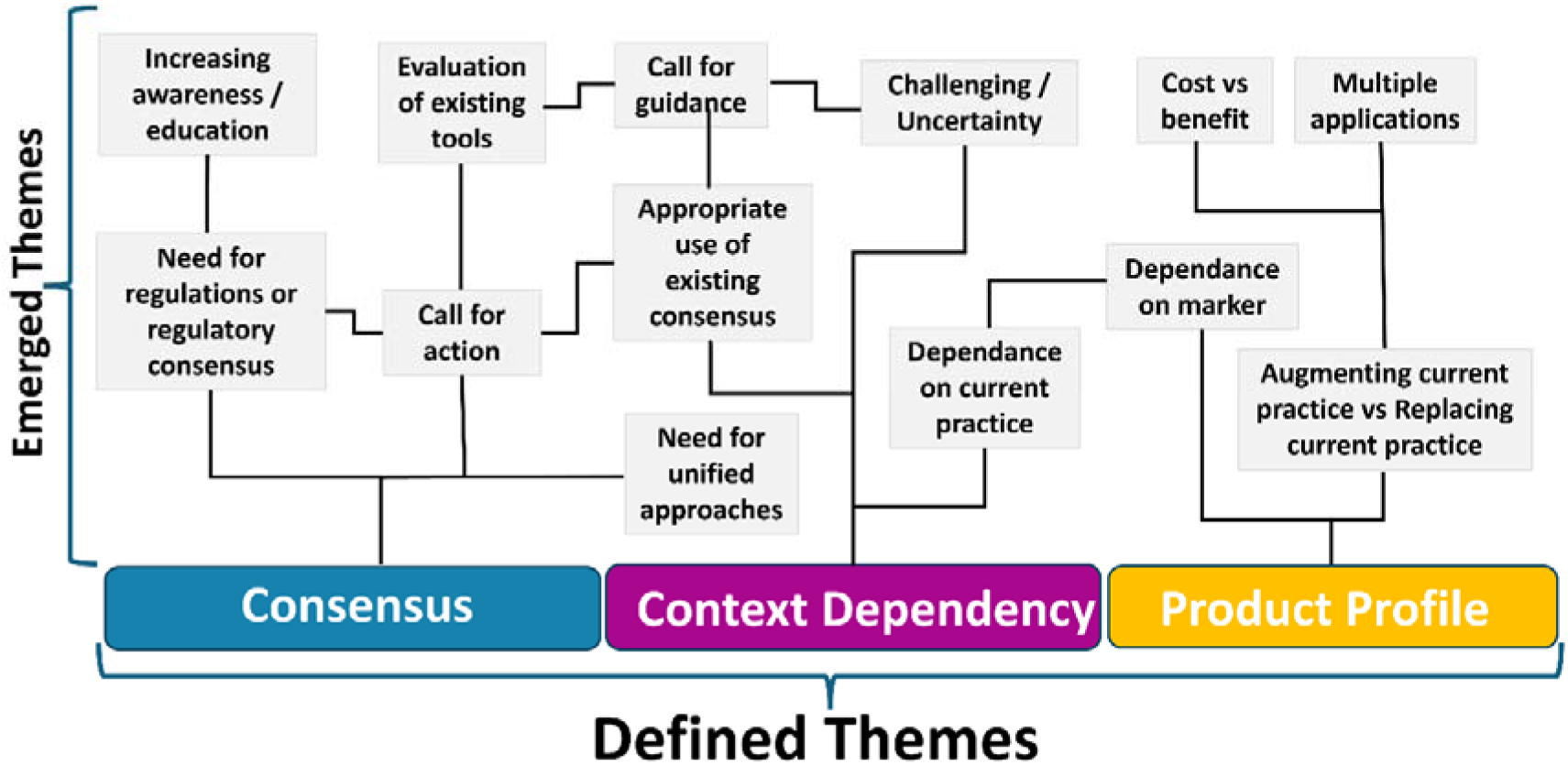
Schematic describing the emerging themes from the free text responses to the survey. From these, the three themes used in subsequent analysis were defined.

Section 4 was based on the question **‘How do we standardise data acquisition and analysis more effectively?’** and Q4A asked ‘…when should we use a phantom? Should we use phantoms to standardise data acquisition for every sequence?’ The results suggested that almost half of the participants (48%) believed it unnecessary, a similar proportion were unsure (43%), whilst only 9% felt it necessary. 52% agreed with a goal-orientated approach to quality (Q4B). A varied response was obtained for Q4C which asked, ‘Should we standardise, harmonise or optimise pulse sequences in multi-centre studies?’, with 42% saying ‘Harmonise’, 24% ‘Standardise’, 24% ‘Unsure’ and 11% ‘Optimise’.

Section 5’s sub-questions came from the question, **‘How do we share data, code, and good practice more effectively?’** 73% thought the code should be shared at the point of publication (Q5Ai) with 71% suggesting that is also when data should be shared (Q5iAi). The remaining section 5 questions were free text responses and consequently no descriptive analysis can be provided. Section 6 discussed Quality Management Systems (QMS), based on the question **‘How can we improve quality management of qMR imaging biomarkers?’** 46% of participants had a QMS at their place of work, but only 50% of those participants used it. The remaining questions in sections 6 and 7 were free-text responses.

### 3.3 Thematic analysis

The 13 emerging themes were colour-coded and re-applied to the whole qualitative dataset to derive all extracts of text that were related. A re-review of the related text extracts developed three broad areas of interest were then defined as Consensus, Context Dependency and Product Profile.

#### 3.3.1 Consensus

The free-text responses revealed that survey participants believe there is a need to build consensus and resources dedicated to improving the standardisation of many aspects of imaging biomarker development, including: (i) terminology, (ii) decision making and (iii) validation. Views were frequently expressed on: (i) the format for sharing consensus, (ii) what consensus building should target, (iii) the lack of multi-disciplinary guidelines, (iv) standardisation of terminology, (v) the need for action groups and (vi) for publications on how to define the translational pathway. These were defined as the theme of Consensus.

#### 3.3.2 Context Dependency

The theme of *Context Dependency* was frequently raised across all topics, including: (i) phantoms, (ii) terminology, (iii) decision making and (iv) endpoints. Views were expressed strongly regarding the uniqueness of each clinical situation when integrating imaging biomarkers into the clinical workflow. The nature of this integration raised some variability in views. The most common perspective suggested the context should be developed based on the imaging biomarkers themselves, closely followed by the clinical question, availability, and cost. The need for the imaging biomarker pathway to be unique may appear diametric to the above expression that there is a need for consensus building. However, examining the two themes together identifies the required nuance to better understand the challenges of clinical translation.

#### 3.3.3 Product Profile

A product profile is a stated set of product characteristics, including intended use, target populations and other desired attributes of products, including safety and efficacy-related characteristics^15^.The theme of *Product Profile* arose across several questions. Opinions strongly favoured qMR imaging biomarkers supplementing, rather than replacing existing pathways. Many considered that a specific endpoint of an imaging biomarker in clinical practice would have a staged approach, although this varied when considering imaging biomarkers across patient groups or along a treatment process. For example, in Q2C, ‘Should change in clinical practice be for all patients or selected cohorts?’, many stated that the ideal situation would be to target clinical translation of an imaging biomarker for a single indication/patient group initially, and then, following a subsequent phased research and development, transfer the imaging biomarker to other patient pathways.

## 4 Discussion

We have conducted the first comprehensive survey of the UK MRI community to understand the barriers and challenges to the clinical translation of qMRI. We have identified 3 main themes: 1) the need to develop *Consensus* in terminology, decision making and validation; 2) an appreciation of uniqueness of each clinical situation and the need for *Context Dependency* ; and 3) the importance of developing a *Product Profile* as early in development as possible. These results highlight the need for the involvement of a range of stakeholders, both in the UK and internationally, and for a framework to be developed to support the translational pathways for qMR imaging and spectroscopic biomarkers to maximise clinical potential.

### 4.1 Consensus

Survey questions such as ‘*In regard to sensitivity and specificity, what percentage of change is significant enough to change practice?* ’ (Q2A) and ‘*Who should be involved in the decision to change practice?* ’ (Q2B) are important, as they address the underlying lack of consistent evidence when assessing the utility of a quantitative imaging biomarker for clinical consumption^16^. The pillar of ‘evidence-based practise’ in areas where there is insufficient or inconsistent data has frequently been supported with Consensus reports^17^. Current consensus papers^18^ generally only include the opinion of experts. It is important to consider the importance of a broader input, in both the experience of those involved and their knowledge base^17^. Depending on the subject matter, a small group of experienced experts may not have multi-disciplinary or multi-profession expertise necessary for practical advice and may also bias the results of the consensus^19^. It is anticipated that the more varied the input, in terms of experience and knowledge, the more useful the information and the greater the chances of obtaining an in-depth consensus that will provide real practical solutions to support the clinical translation qMR imaging biomarkers.

Question 2B, ‘*Who should be involved in the decision to change practice?* ’, demonstrated that it was expected that translating an imaging biomarker would require a breadth of input from the MRI community, with 88% of participants indicating that 2 or more groups should be involved in decision making. Participant 6 stated the need for the “*Widest possible range of international stakeholders e.g., radiologists, physicists, device manufacturers, pharmacokinetics/modellers, regulators* .” Participant 7 stated that it is important to “*cast a very wide net here, involve all stakeholders early, build trust and prepare ground for change. These are established knowledge sharing principles* ”. The adoption of a flexible framework for consensus building, which allows a wide range of opinions to affect the outcome would be favourable. This is supported by a comment by participant 62, that “*All should be involved, but more important to define the role/impact each person has in managing/affecting[sic] the change* ”. However, a multi-disciplinary approach must be weighed against the practicalities and complexity of completing and publishing a consensus in an achievable timeframe.

Question 1B demonstrated over two-thirds of participants wanted resources to promote the use of standardised language in consensus papers. Standardising language can be a useful tool in improving effective communication^20^ in complex situations, such as developing quantitative imaging biomarkers, the intrusion of jargon or differentiating lexicons from a network of professions means we are at risk of miscommunication. A point noted by participant 57 was that “…*manufacturers have their own terminology* ”. Participant 31 suggested a solution, the use of a “…*a dictionary for reporting scan data and pre-processing steps, e.g., by OSIPI* ^21^.” Participant 5 also stated that we need “*Consensus building of agreed lexicons and shared glossary for each qMR method, e.g., IPEM SIG publish recommended language and precise definitions.* ”

In addition to shared language, shared code and data were also seen as important. Question 3C showed that participants strongly agreed that institutions needed to provide knowledge support. However, participants agreed that both funders and publishers should mandate sharing of both code and data, with participant 29 stating that “*Open Sharing should be the norm* .”^22^

### 4.2 Context Dependency

Adopting a consensus approach allows the qMR imaging biomarker communities to express the importance of context dependency in a range of areas, such as technical specifications and acceptable diagnostic quality. However, the methodology used to reach consensus is a possible area of improvement. An example of this is the ASL community, who in 2015 developed recommendations for implementing ASL for clinical application.^23^ This was performed by two working groups running 2 consecutive workshops in a single location. Later, in 2019, further recommendations for clinical translating renal ASL^24^ were developed. This was performed by a panel of international experts, following the adopted Delphi method using on-line surveys and in-person meetings. It could be argued that this more structured and holistic approach reduced bias and increase completeness. Recent consensus building has developed guidance for a range of imaging biomarkers in broader contexts^1,2^, nevertheless consensus methodology is highly variable, and more work is needed to establish the more standardised, adaptive approach that was suggested by the survey results. Question 1B allowed a free text response to ‘*Should effort be made to develop a consensus paper to standardise terminology to aid clinical translation?* ’ and over 70% of participants stated that there should be. Participant 36 noted that “*Consensus papers are really important, but they should relate to discrete domains/focussed on organs and/or disease processes.* ”

Q2A, ‘*In regard to sensitivity and specificity, what percentage of change is significant enough to change practice?* ’, had a more even distribution of responses, also reflected in follow-up remarks than other survey questions. This diversity of opinion is likely due to the required sensitivity and specificity being context dependent and the participants considering their answers with respect to their own area of expertise. Similarly, Q1C stated that ‘*Quantitative MR parameter maps often need to be created away from the scanner console* ’ and asked, ‘*What is the maximum amount of time after acquisition, if an imaging biomarker is to be integrated into the clinical workflow?* ’, results were variable with participant 45 commenting that “…*Diagnostic and treatment pathways vary enormously across clinical practice, and the urgency for diagnostic information varies too. I find it difficult to see the value of this question unless the intended purpose is focused on a particular scenario..* .” and participant 40 saying, “*This is highly dependent on the urgency of the indication, and all 4 options are possible depending on the context.* ” Similarly, responses were varied for Q4C which asked ‘*Should we standardise, harmonise or optimise pulse sequences in multi-centre studies?* ’. Again participant 45 commented “*These questions don’t capture the broad spectrum of sequences, research questions, and requirements of the many different types of multisite study. I don’t think you can give yes/no answer or a rigid process/definition for validate/standardise/harmonise/optimise that fits the breadth of activities covered by biomarker development and application* .”

*Context Dependency* is key to developing consensus within useful and workable boundaries. How we decide on those boundaries is an interesting and open question and one that requires further discussion.

### 4.3 Product Profile

Defining *Product Profile* involves stating clearly and unambiguously the intended use of a qMR imaging biomarker. For instance, the intended medical indication, patient population, or body part, as well as the intended technical environment and where in the clinical pathway it will fit. This mirrors the process for the development of Software as a Medical Device. Integration into the clinical pathway via the Picture Archiving and Communication System (PACS), which is used to securely store and digitally transmit electronic images, was address in Q7B ‘*How do we integrate our MR development with PACs systems?* ’ Participant 12 stated the obvious need to “*To engage with people that involve in developing and maintaining PACS system* ” and participant 18 highlighted the need for “*better coordination between NHS Trusts and Universities to iron out the potential data safety issues* .” Participant 33 suggested that we “*Specify “research access” at time of PACs tender* .”

A clear *Product Profile* can aid the research design and end translation of a medical test ^25,26^, but more input from the MRI community would be required to establish clear product profiles for individual qMR imaging biomarkers. Participants often stated conversations required increased input from a larger breadth of stakeholders and networks, such as vendors and national/international societies involved in quantitative MR. Dialogue with patients is also key, a subject touched upon in Q7C, ‘*How do we find out if a patient group can tolerate the imaging method?* ’. Many supported that inclusion of Patient and Public Involvement (PPI) early in study and the early feasibility studies on patients with participant 4 stating with need to “*Engage with PPI groups and run small bespoke patient studies before starting a larger study.* ”

There is an acknowledged reticence towards the use of phantoms by the manufacturers and Q4A was prefaced with sentiment gained from the roundtable discussion that ‘It is known that vendors will find a method less attractive if a method requires a phantom’. The results suggested that almost half of the participants (48%) believed it unnecessary to use a phantom for every sequence. As stated in Question 5C, ‘*Techniques generally require buy-in from scanner manufacturers to become part of their product in order to change clinical practice* .’ Therefore, the view of the manufacturers can both influence and curtail the ability of researchers to engage in practices that are believed to improve imaging biomarker reproducibility ^27–30^

The World Health Organization (WHO) has adopted Target Product Profiles (TPPs). TPPs define the required characteristics focused to the aimed disease. They consider the intended use, target populations, attributes that will ensure safety, efficacy and feasibility. By establishing the TPP at an early stage can provided needed guidance during development and ensure that emerging techniques are meeting all unmet clinical needs^25^. Group 7 focused on engaging clinicians to support the translation process with the question *‘How can we engage with end users to support clinical translation?* **’** (Q7A) and developed questions that addressed the need to be guided by the end result. Although the theme here spoke more broadly than the development and distribution of formal TTPs, all responses gave insight into improving meaningful discussion between all levels of the translational pipeline to keep development focused on the unmet clinical need.

### 4.4 Limitations

Although the questions asked in this survey were derived from a broad-ranging roundtable discussion, that was preceded by a comprehensive overview of the problems and potential solutions to the challenges of qMR biomarker translation, there are some limitations.

Firstly, the seed questions for the 7 sections debated at the roundtable discussion were chosen by a multi-disciplinary team of BIC-ISMRM workshop co-organisers with significant collective experience in qMR methods development, use and clinical practice (PC, KM, HH). Although designed to focus on areas pertinent to clinical translation and encourage meaningful conversation, they will have heavily influenced the discussion and subsequently the survey questions that evolved. Similarly, co-authors (PC, JM) were involved in formatting the survey question wording and the possible answers/answer types.

Secondly, the round-table discussion was at the BIC-ISMRM conference, which is most popular with research MR physicists. Although, it is important to note that all workshop attendees had heard a wide range of perspectives in the preceding workshop talks. The survey was sent to several mailing lists, as well as to the conference attendees, however 64% of the responses were from Research Imaging Scientists.

Lastly, a priority to build, test and release the survey whilst attendees were still present at the conference was important to maximise the response rate. We acknowledge that the methodology to review the wording of questions could have been improved. For further work, we will consider using a focus group composed of a broad range of specialities to review proposed questions ahead of distribution.

### 4.5 Implications and future work

The admittance that the translational pipeline is ineffective is not unique to qMR or the UK. In 2008 the Swiss Academy of Medical Sciences “SAMS” referred to the gaps faced in translating research as the “valley of death” ^31^. In the US, the same year, Simon^32^ discussed the oncology treatment research noting serious limiting factors that created discrepancies between the scientific discoveries and the translational clinical interventions. More recently, Ioannidis et al ^33^ described the biomarker pipeline as wasteful and with “leaks”, concluding that the attempt at clinical translation is too often synonymous with failure. With further comment that when reflecting on the causation of these challenges can be grouped by those faced by the biomarker community as a whole and challenges that are individual to the “peculiarities” of the niche research landscape, such as qMR imaging biomarkers.

Future work looks to extend this discussion to other stakeholders in the clinical translation of qMR imaging biomarkers. This could be done by holding similar workshops at both clinical and radiographer focused scientific meetings, for instance European Congress of Radiology and conferences hosted by the Society of Radiographers, and by extending our research and framework internationally. This outreach has already started with a member-initiated symposium at the International Society of Magnetic Resonance in Medicine meeting held in Singapore in 2024. Attendees heard the perspectives of 5 key international voices in qMR biomarker translation and had the opportunity to participate in a live, interactive survey throughout the symposium.

The researchers involved in this process continue to develop the STEPS initiative with the following mission:

> “To facilitate the translation of quantitative magnetic resonance imaging and spectroscopy biomarkers from research to clinical practice by promoting collaboration between researchers, technologists, industry, regulatory bodies, and stakeholders including patients, carers and the public. Through open communication and shared knowledge, we aim to establish a framework for the development and validation of magnetic resonance imaging and spectroscopy biomarkers that meet the needs of both patients and healthcare providers. By prioritizing transparency, ethical standards, and scientific rigor, we strive to build trust and confidence in these novel quantitative imaging and spectroscopy biomarkers in clinical settings, ultimately improving patient outcomes and transforming healthcare.”

By developing a ***network-of-networks*** of the key stakeholders in qMR biomarker translation within the UK, recognising the obstacles along this pipeline, and showcasing the fundamental studies and initiatives that are already improving the translational landscape, it is hoped that STEPS can be a framework of support that empowers knowledge sharing and creates resources to support the delivery of personalised, non-invasive, game-changing, quantitative imaging biomarkers to the clinic.

## 4. Conclusions

The survey of the UK MRI community on the clinical translation of qMR imaging and spectroscopic biomarkers discovered three main themes: 1) the need to develop *Consensus* in terminology, decision making and validation; 2) variability in the responses due to uniqueness of each clinical situation highlighted the requirement for *Context Dependency* in the process of consensus building; and 3) the importance of stating clearly and unambiguously the intended use of the biomarker and developing a *Product Profile* .

The next steps involve broadening this conversation to involve more stakeholders, both in UK and internationally, and developing a framework to support the translational pathways for qMR imaging and spectroscopic biomarkers from pre-clinical techniques to research methods and eventually effective clinical decision-making tools.

## Data Availability

All data produced in the present study are available upon reasonable request to the authors.

